# Identifying Louisiana communities at the crossroads of environmental and social vulnerability, COVID-19, and asthma

**DOI:** 10.1101/2021.07.19.21257742

**Authors:** Arundhati Bakshi, Alicia Van Doren, Colette Maser, Kathleen Aubin, Collette Stewart, Shannon Soileau, Kate Friedman, Alexis Williams

## Abstract

The COVID-19 pandemic has disproportionately affected the socially and environmentally vulnerable, including through indirect effects on other health conditions. Asthma is one such condition, which may be exacerbated by both prolonged adverse in-home exposures if quarantining in unhealthy homes and prolonged outdoor exposures if the ambient air quality is unhealthy or hazardous. As both are often the case in Environmental Justice (EJ) communities, here we have analyzed data at the census tract (CT) level for Louisiana to assess any correlation between social and environmental vulnerability, and health issues like COVID-19 and asthma. Higher Social Vulnerability Index (SVI), Particulate Matter less than 2.5 µm in diameter (PM_2.5_) and Ozone levels were associated with higher rates of cumulative COVID-19 incidence at various time points during the pandemic, as well as higher average annual asthma hospitalization rates and estimated asthma prevalence. Further, cumulative COVID-19 incidence during the first three months of the pandemic was moderately correlated with both asthma hospitalizations and estimated prevalence, suggesting similar underlying factors may be affecting both conditions. Additionally, 137 CTs were identified where social and environmental vulnerabilities co-existed, of which 75 (55%) had high estimated prevalence of asthma. These areas are likely to benefit from asthma outreach that considers both social and environmental risk factors. Fifteen out of the 137 CTs (11%) not only had higher estimated prevalence of asthma but also a high burden of COVID-19. Further research in these areas may help to elucidate any common social determinants of health that underlie both asthma and COVID-19 burdens, as well as better clarify the possible role of the environment as related to the COVID-19 burden in Louisiana.

## Introduction

In 2020, the COVID-19 pandemic laid bare the major health impacts of social and environmental vulnerability, by taking a heavy toll on populations suffering such vulnerabilities. It highlighted how a novel virus does not affect everyone equally; it exploits underlying health conditions that are more prevalent in vulnerable populations [1, 2]. According to the US Centers for Disease Control and Prevention (CDC), asthma is one such respiratory health condition that might be a risk factor for severe illness with COVID-19. It is not only more prevalent among African-Americans and people living in poverty (a population disproportionately affected by COVID-19 [3, 4]), but asthma is also heavily influenced by environmental factors [5-7]. As such, it is particularly important to tackle this issue in Environmental Justice (EJ) communities, which lack equitable access to protection from environmental and health hazards, and where socioeconomic, demographic and environmental stressors overlap to create environmental health disparities. For example, many people living in EJ communities do not have access to affordable healthy housing, and thus may be exposed daily to asthma triggers in the home [8]. At a time when people are spending more time in their homes than ever, many asthmatics living in unhealthy homes may experience worsening of their symptoms. People living in communities that have outdoor air concerns may experience prolonged adverse outdoor exposures to known asthma triggers, such as ozone and Particulate Matter less than 2.5 µm in diameter (PM_2.5_), if they spend more time socializing outdoors in keeping with COVID-19 safety protocols. These, in turn, could translate into poor outcomes if they contracted COVID-19.

The need to tackle this vicious cycle was recently highlighted by a study that showed respiratory health risks from hazardous air pollutants and high incidence of COVID-19 converged in several counties across the southeastern United States [9]. These areas tended to have higher non-Hispanic black populations, socioeconomically disadvantaged persons, people with disabilities and people without health insurance. This county-level study served as an important starting point for further exploring the impact of COVID-19 in EJ communities at a finer geographic resolution on a state-by-state basis [9]. Since Louisiana (LA) was one of the states where a majority of counties (called ‘parishes’ in LA) seemed to fall under the “high-high” category for both respiratory hazard risk and COVID-19 risk, here we have undertaken a census tract (CT)-level analysis to assess any correlation between social and environmental vulnerability, COVID-19, and asthma. We have further identified several EJ communities with a high burden of asthma and COVID-19 where mitigation measures may be targeted, as well as where further studies may be conducted to elucidate the environment’s role on COVID-19 outcomes in Louisiana.

## Methods

### Indicators of social vulnerability

To assess social vulnerability, we used the 2018 Social Vulnerability Index (SVI) derived by the CDC Agency for Toxic Substances and Disease Registry (ATSDR) [10]. Along with the Overall SVI, we considered the four specific SVI themes outlined by CDC/ATSDR – socioeconomic status (Theme 1); household composition and disability (Theme 2); minority status and language (Theme 3); and housing type and transportation (Theme 4). We also applied the US Census Bureau’s 2018 American Community Survey (ACS) data for percent population without health insurance, below poverty level, unemployed, without a high school diploma, disabled, minority (all persons except non-Hispanic white), age 65+, age 17 and under, and population (age 5+) who speak English “less than well”. We also took into account household level information on those without access to a vehicle, single parent households and households with more people than rooms (crowded homes). Detailed information on each of these variables is available from CDC/ATSDR (https://www.atsdr.cdc.gov/placeandhealth/svi/documentation/SVI_documentation_2018.html).

All datasets for social vulnerability **(Table 1)** were considered at the census tract level, and the SVI scores classified into five categories based on the CT’s ranking (percentile) against all CTs in the state: 0-0.20; 0.21-0.40; 0.41-0.60; 0.61-0.80; 0.81-1.00. All mapping and spatial analyses were performed in ESRI ArcGIS 10.3.1.

**Table 1.** Summary of datasets assessed in this study. All datasets were mapped at the census tract level.

| Topic | Indicator (Measure) | Data Source | Data Year(s) |
| --- | --- | --- | --- |
| Social Vulnerability | Social Vulnerability Index and related indicators from ACS 2018 | CDC/ATSDR | 2018 |
| Environmental Vulnerability | NATA Respiratory Hazard Index | EPA | 2014 |
|  | PM<sub>2.5</sub> level (average annual concentration in µg/m<sup>3</sup>) | EPA | 2016 |
|  | Ozone level (summer seasonal average of daily maximum 8-hour concentration in air in parts per billion) | EPA | 2016 |
|  | Indoor mold concerns reported to IEQES program (average annual number of calls) | LDH | 2017-2019 |
| COVID-19 | Cumulative COVID-19 incidence rate at 3-, 6-, 9- and 12-month time points | LDH | Mar 2020-Mar 2021 |
| Asthma | Asthma Hospitalization (average annual crude rate, where asthma was a primary diagnosis among hospitalization cases) | LDH | 2017-2019 |
|  | Estimated Asthma Prevalence (average annual crude rate, where asthma was any diagnosis among hospitalization cases) | LDH | 2017-2019 |

### Indicators of environmental vulnerability

In terms of environmental datasets (outdoor air), we downloaded the following from US Environmental Protection Agency’s (EPA’s) EJSCREEN tool version 2020 (https://www.epa.gov/ejscreen): National Air Toxics Assessment (NATA) Respiratory Hazard Index (RHI), PM_2.5_ levels and Ozone levels. The EJSCREEN technical documentation (available from: https://www.epa.gov/sites/production/files/2021-04/documents/ejscreen_technical_document.pdf) provides detailed definitions for each of these measures. Briefly: The RHI is the ratio of the exposure concentration of Hazardous Air Pollutants (HAPs) to a health-based reference concentration. The most recent year of RHI data available (2014) takes into account 181 HAPs (including diesel particulate matter), for which both emissions and health-related toxicity data were available. In order to derive the RHI, US EPA uses several modeling techniques to estimate both the concentration of the HAPs as well as the risk of exposure and downstream respiratory health risks. Given that the susceptibility to air pollution cannot be reliably estimated, these models take into consideration differences in emissions and meteorology (which affects the concentration of pollutants in ambient air) and the differences in the location of individuals (which affects exposure) among CTs. Thus, applying these models then allow us to identify geographic patterns and assess relative risks at the CT-level, with the caveat that all calculations become more uncertain with smaller geographies. Complete details on RHI modeling and calculation methods are available from the 2014 NATA Technical Support Document [11]. PM_2.5_ levels refer to average annual concentrations in µg/m^3^, as calculated by US EPA using a fusion of modeled and monitored data (for methodological details on air quality modeling, see [12]). Ozone levels were similarly calculated using a combination of modeled and monitored data by the EPA [12], using the summer seasonal average of daily maximum 8-hour concentration in air in parts per billion. The most recent data available for both PM_2.5_ and Ozone levels (2016) was used for this study.

Finally, to account for not only outdoor but also Indoor Environmental Quality (IEQ) concerns, we surveyed internal data from the Louisiana Department of Health’s (LDH’s) Indoor Environmental Quality Education Service (IEQES). Since 2000, LDH’s IEQES has been providing guidance to Louisiana homeowners and renters on a variety of IEQ concerns reported to the program. The majority of these calls (∼75%) are related to mold in residential settings; however, the IEQES program provides guidance on all types of IEQ issues (including odors, pests, radon, chemical exposures, etc.) in both private and public buildings (such as homes, schools, places of worship, etc.). For this study, we have aggregated the three most recent years of data available through IEQES (2017-2019) and calculated the average annual number of mold concerns reported to the program as an indicator of IEQ concerns in an area. Mold concerns were chosen for this study over all IEQ concerns since they comprised the majority (86%) of the calls received. Further, mold concerns were most often associated with respiratory complaints (the health focus of this study), as reported by the callers; of the 785 calls that reported respiratory complaints, 708 (90%) were associated with mold. Mold exposure is also recognized as a possible environmental trigger for asthma [13].

All environmental datasets **(Table 1)** were considered at the census tract level. For all datasets, we used EJSCREEN’s visualization method and mapped the RHI, PM_2.5_, Ozone and IEQ concerns as a percentile score relative to all CTs in the state (ranked using the PERCENTRANK.INC function in MS Excel 2016). A choropleth map was then created using progressively darker colors for five categories: 0-20^th^ percentile; 21^st^-40^th^ percentile; 41^st^-60^th^ percentile; 61^st^-80^th^ percentile and 81^st^-100^th^ percentile.

### Health indicators (COVID-19 and asthma)

Total number of COVID-19 cases by CT were downloaded from LDH’s COVID-19 dashboard (https://ldh.la.gov/coronavirus/) and cumulative incidence rates calculated at four time points (3, 6, 9 and 12 months into the pandemic in LA) by dividing the total number of cases at each time point by the ACS 2018 population of the CT. Of the 1145 CTs in LA, data were available for approximately 1034 CTs at the 3-month time point, 1038 at the 6-month time point, and 1039 at the 9- and 12-month time points. Case counts are not provided for CTs with a 2018 population of less than 800 to safeguard privacy.

Since asthma surveillance data were not available at the census tract level for most of Louisiana, we estimated asthma burden using the inpatient discharge data available through LDH. To minimize the need for suppression, inpatient discharge data was aggregated for the three most recent years available (2017-2019) and average annual crude rates were calculated for cases where asthma (ICD-10 code J45) was the primary diagnosis, as well as where asthma was any diagnosis. While the former indicated the level of asthma exacerbation in a community that required medical intervention (an indicator for uncontrolled asthma), the latter served as an estimate of asthma prevalence in the community. In order to calculate these, we divided the total number of cases for each CT (2017-2019) by the total population of the CT during that period, and then divided that value by 3 to yield the average annual crude rates.

Similar to the environmental vulnerability datasets, percentile scores were also calculated for COVID-19 incidence and asthma data **(Table 1)**, which were then classified in five categories.

### Correlation analysis

Spearman’s Rank Correlation was utilized to analyze the correlation between various social and environmental vulnerability factors, COVID-19 incidence, and the measures of asthma risk by CT. This was performed by first ranking the values in each dataset using RANK.AVG function in MS Excel 2016, followed by applying the PEARSON function to compare two datasets. Significance was set at alpha less than 0.05 (α < 0.05), with degrees of freedom (df) equal to two less than the total number of data points represented in both datasets. Only CTs for which all data were available (1035 out of 1145 total CTs in LA) were used for this analysis. Spearman’s *rho* (ρ) was used to identify the direction and strength of correlation. If the p-value was less than 0.05 (p < 0.05), strength of correlation was defined as follows: absolute value of ρ = 0.01-0.30 = weakly correlated; 0.31-0.70 = moderately correlated; and 0.71-1.00 = strongly correlated. The direction of correlation was determined by the sign (positive or negative) on ρ value.

## Results

### Relationship between social and environmental vulnerability, COVID-19, and asthma

CTs were defined as ‘high’ for social vulnerability if the Overall SVI was greater than or equal to 0.75, and ‘high’ for environmental vulnerability if the RHI, PM_2.5_ level or Ozone level was at or over the 75^th^ percentile for all CTs in the state. When compared to all CTs, high SVI was associated with higher average cumulative COVID-19 incidence rates early in the pandemic (3-month and 6-month time points). PM_2.5_ was associated with lower COVID-19 incidence rates at the 3-month time point, but at 12 months, it was associated with higher COVID-19 incidence. Ozone levels were associated with higher COVID-19 incidence all time points (except at 6 months), whereas RHI showed the opposite pattern, being associated with significantly lower COVID-19 incidence at all time points except at 6 months **(Fig 1A)**. Additionally, the COVID-19 incidence rates were higher in areas of high SVI + high Ozone compared to areas with only high SVI. This additive effect over SVI was not observed from any other environmental factor, indicating that Ozone may be an important environmental factor affected COVID-19 incidence. With regard to average hospitalization rate for asthma and estimated asthma prevalence, higher rates of both were associated with high SVI and high Ozone levels **(Fig 1B)**. High SVI appeared to have an additive effect on high Ozone, with areas of high SVI + high Ozone having higher asthma hospitalization rate and estimated prevalence than areas that just had high Ozone. Taken together, these descriptive statistics indicated a possible relationship between social and environmental vulnerability (especially related to Ozone), COVID-19, and asthma.

**Figure 1.**
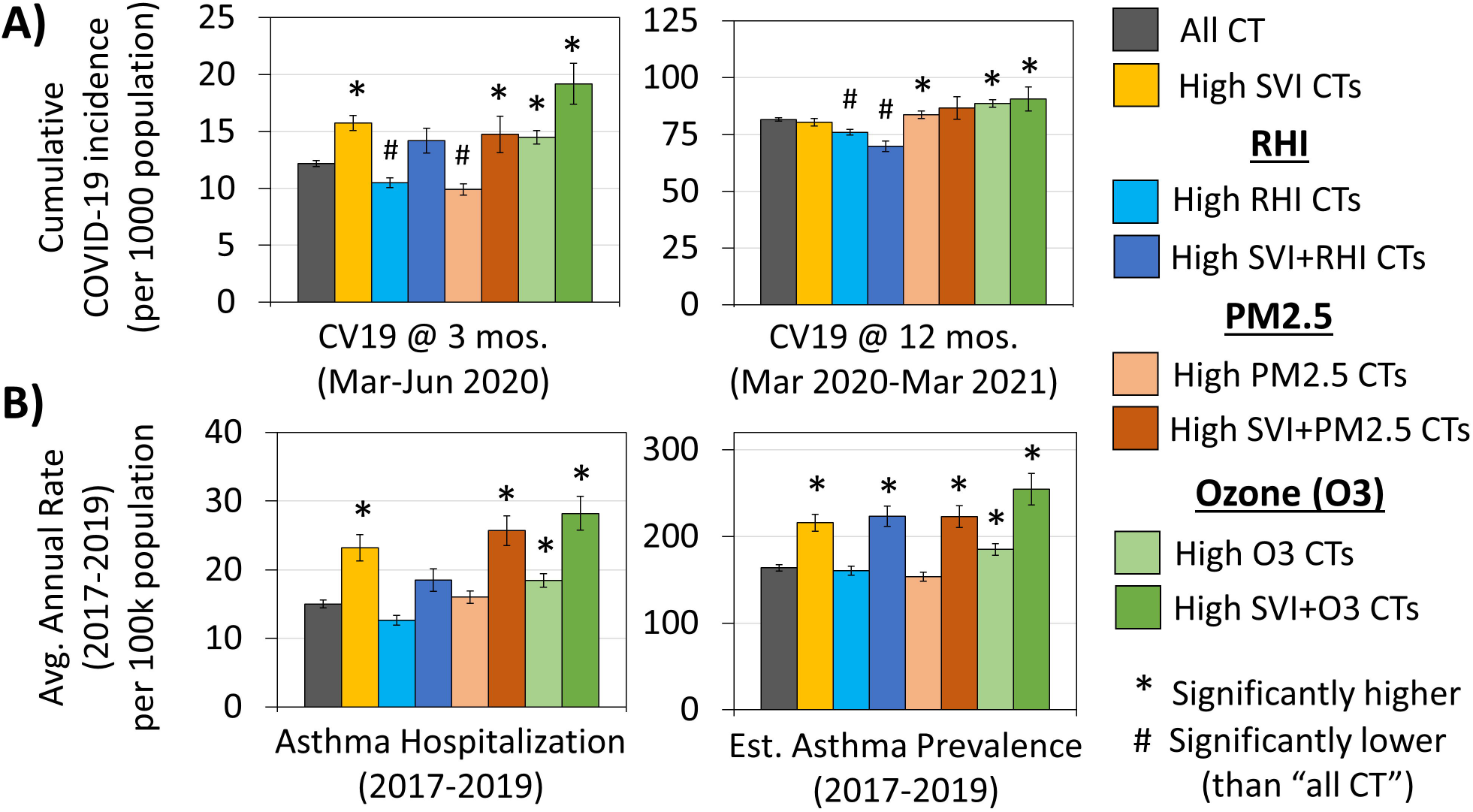
COVID-19 and Asthma in areas with high social and/or environmental vulnerability. **(A)** High SVI was associated with higher average cumulative COVID-19 incidence rates early in the pandemic, whereas PM_2.5_ was associated with higher rates later in the pandemic. Cumulative COVID-19 incidence was elevated in areas of high Ozone at all points during the pandemic. **(B)** Asthma hospitalization as well as estimated asthma prevalence followed a similar pattern with higher rates of both being associated with high SVI and high Ozone. Error bars indicate 95% confidence intervals; asterisks (*) indicate areas that are significantly higher than “All CTs” and pounds (#) indicate areas significantly lower than “All CTs” (p<0.05; T-test).

These possible relationships were then further explored in a Spearman rank correlation **(Table 2 and 3)**. Cumulative COVID-19 incidence rate at the 3-month time point was moderately correlated with the Ozone levels (ρ=0.32; p<0.0001), SVI for minority status and language barriers (ρ=0.44; p<0.0001), asthma hospitalization (ρ=0.32; p<0.0001), and estimated prevalence of asthma (ρ=0.50; p<0.0001). Asthma hospitalization and estimated prevalence were moderately correlated with SVI for socioeconomic status, SVI for minority and language barriers and Overall SVI (ρ=0.30-0.37; p<0.0001). Since one of the strongest correlation coefficients was observed between the estimated prevalence of asthma and cumulative COVID-19 incidence at the 3-month time point, we conducted a second Spearman rank correlation to test whether similar social vulnerability measures may underlie both health indicators **(Table 3)**. According to the results, positive correlations of moderate strength were observed between both health indicators and percent minority population (ρ=0.51-0.52; p<0.0001) and percent households without a vehicle (ρ=0.37-0.44; p<0.0001).

**Table 2.** Spearman’s rho (correlation coefficient) for social and environmental vulnerability, asthma, and COVID-19. With regards to COVID-19, strongest correlation was observed between the cumulative incidence during the first quarter of the pandemic in LA (Mar-Jun 2020) and the estimated prevalence of asthma, SVI due to minority status and language barriers, asthma hospitalization crude rate, and the percentile score for ozone levels. With regards to asthma, SVI due to socioeconomic status, minority status and language barriers, and overall SVI were also significantly correlated with both asthma hospitalization rate as well as the estimated prevalence.

|  |  | Hosp.<br>Asthma | EPV<br>Asthma | CV19<br>(3m) | CV19<br>(6m) | CV19<br>(9m) | CV19<br>(12m) |
| --- | --- | --- | --- | --- | --- | --- | --- |
| Environmental<br>Factors | RHI | -0.02 | 0.04 | -0.12 | -0.05 | -0.14 | -0.14 |
|  | Ozone | 0.21 | 0.29 | 0.32 | 0.03 | 0.09 | 0.11 |
|  | PM2.5 | 0.08 | 0.02 | -0.07 | -0.02 | 0.12 | 0.08 |
| Social<br>Vulnerability | SVI_SES | 0.36 | 0.34 | 0.22 | 0.15 | -0.16 | -0.24 |
|  | SVI_HC | 0.26 | 0.26 | 0.09 | 0.16 | 0.01 | -0.02 |
|  | SVI_ML | 0.30 | 0.37 | 0.44 | 0.29 | -0.01 | -0.06 |
|  | SVI_HT | 0.13 | 0.09 | 0.11 | 0.19 | 0.02 | -0.07 |
|  | SVI_OV | 0.37 | 0.36 | 0.27 | 0.26 | -0.06 | -0.15 |
| Asthma | Hosp. Asthma |  | 0.61 | 0.32 | 0.21 | 0.02 | 0.01 |
|  | EPV Asthma | 0.61 |  | 0.50 | 0.28 | 0.03 | 0.07 |
| COVID-19<br>(cumulative) | CV19 (3m) | 0.32 | 0.50 |  |  |  |  |
|  | CV19 (6m) | 0.21 | 0.28 |  |  |  |  |
|  | CV19 (9m) | 0.02 | 0.03 |  |  |  |  |
|  | CV19 (12m) | 0.01 | 0.07 |  |  |  |  |
| Abbreviations: RHI = NATA Respiratory Hazard Index; Ozone = Ozone levels; PM2.5 = PM<sub>2,5</sub> levels; SVI_SOCEC = Social Vulnerability Index (due to socioeconomic factors); SVI_HC = Social Vulnerability Index (due to household composition); SVI_ML = Social Vulnerability Index (due to minority status and language barriers); SVI_HT = Social Vulnerability Index (due to housing and transportation factors); SVI_OV = overall Social Vulnerability Index; Hosp. Asthma = Asthma hospitalization crude rate; EPV Asthma = Estimated prevalence of asthma; CV19 = Cumulative COVID-19 incidence at 3, 6, 9 and 12 months |  |  |  |  |  |  |  |
| Legend based on rho value: |  | Strength and direction of correlation: |  |  |  |  |  |
| >0.7 to 1.0 |  | Strongly positive correlation |  |  |  |  |  |
| >0.3 to 0.7 |  | Moderately positive correlation |  |  |  |  |  |
| >0 to 0.3 |  | Weakly positive correlation |  |  |  |  |  |
| Rho=0 or p ≥ 0.05 |  | No significant correlation |  |  |  |  |  |
| <0 to -0.3 |  | Weakly negative correlation |  |  |  |  |  |
| < -0.3 to -0.7 |  | Moderately negative correlation |  |  |  |  |  |
| < -0.7 to -1.0 |  | Strongly negative correlation |  |  |  |  |  |

**Table 3.** Spearman’s rho (correlation coefficient) for specific social vulnerability factors, asthma and COVID-19. Statistically significant correlations were observed between several vulnerability factors and cumulative COVID-19 incidence up to June 2020 (CV19 [3m]) and/or estimated prevalence of asthma (EPV Asthma). However, the factors that showed moderately strong correlation coefficients with both health indicators were percent minority populations and percent households without a vehicle.

|  | **EPV**<br>**Asthma** | **CV19**<br>**(3m)** | *Abbreviations:*<br>PP/H/U = Percent Population/Household/Housing Unit |
| --- | --- | --- | --- |
| **PP_AGE17** | 0.01 | -0.06 | AGE17 = Age 17 and under; |
| **PP_UNINSUR** | 0.23 | 0.18 | UNINSUR = No health insurance; |
| **PP_POV** | **0.31** | 0.23 | POV = Below poverty; |
| **PP_UNEMP** | 0.29 | 0.16 | UNEMP = Unemployed; |
| **PP_NOHSDP** | 0.28 | 0.17 | NOHSDP = No high school diploma; |
| **PP_AGE65** | -0.01 | 0.01 | AGE65 = Age 65 and over; |
| **PP_DISABL** | 0.29 | 0.05 | DISABL = Disabled; |
| **PP_MINRTY** | **0.51** | **0.52** | MINRTY = Minority; |
| **PP_LIMENG** | 0.10 | 0.18 | LIMENG = Limited English; |
| **PH_SNGPNT** | 0.27 | 0.20 | SNGPNT = Single parent households; |
| **PU_CROWD** | 0.06 | 0.01 | CROWD = Crowded housing units; |
| **PH_NOVEH** | **0.44** | **0.37** | NOVEH = Households without vehicle; |
| **PP_GROUPQ** | -0.06 | 0.07 | GROUPQ = Living in group quarter; |
| *Legend based on rho value:* |  |  | *Strength and direction of correlation:* |
| **>0.7 to 1.0** |  |  | Strongly positive correlation |
| **>0.3 to 0.7** |  |  | Moderately positive correlation |
| **>0 to 0.3** |  |  | Weakly positive correlation |
| *Rho=0 or p ≥ 0.05* |  |  | No significant correlation |
| **<0 to -0.3** |  |  | Weakly negative correlation |
| **< -0.3 to -0.7** |  |  | Moderately negative correlation |
| **< -0.7 to -1.0** |  |  | Strongly negative correlation |

### Identifying EJ communities of concern with respect to COVID-19 and asthma

Surveying the spatial distribution of statewide social vulnerability data showed that while most LA parishes (59/64) had one or more CTs with high overall SVI, i.e. SVI>=0.75 **(Fig 2A)**, environmental vulnerability factors were more likely to be high (at or above the 75^th^ percentile) in certain specific parts of the state. Ozone levels, for instance, were higher in the north, northwest and southeast **(Fig 2B)**; PM_2.5_ levels were higher in the north, northwest and east **(Fig 2C)**; and RHI was higher in the northwest, southwest and eastern parts of the state (map not shown). In terms of IEQ issues, at least one mold concern was reported from 542 CTs in LA (**Fig 2D**), 301 of which (55%) also had high RHI, high PM_2.5_ or high Ozone levels. These CTs, where both indoor and outdoor environments may be of concern, were present in pockets but distributed across the state. A similar inspection of the health measures (asthma and COVID-19) revealed that they were most often noted as ‘high’ (at or above the 75^th^ percentile for the state) in northwestern and southeastern parts of the state **(Fig 2E-F)**.

**Figure 2.**
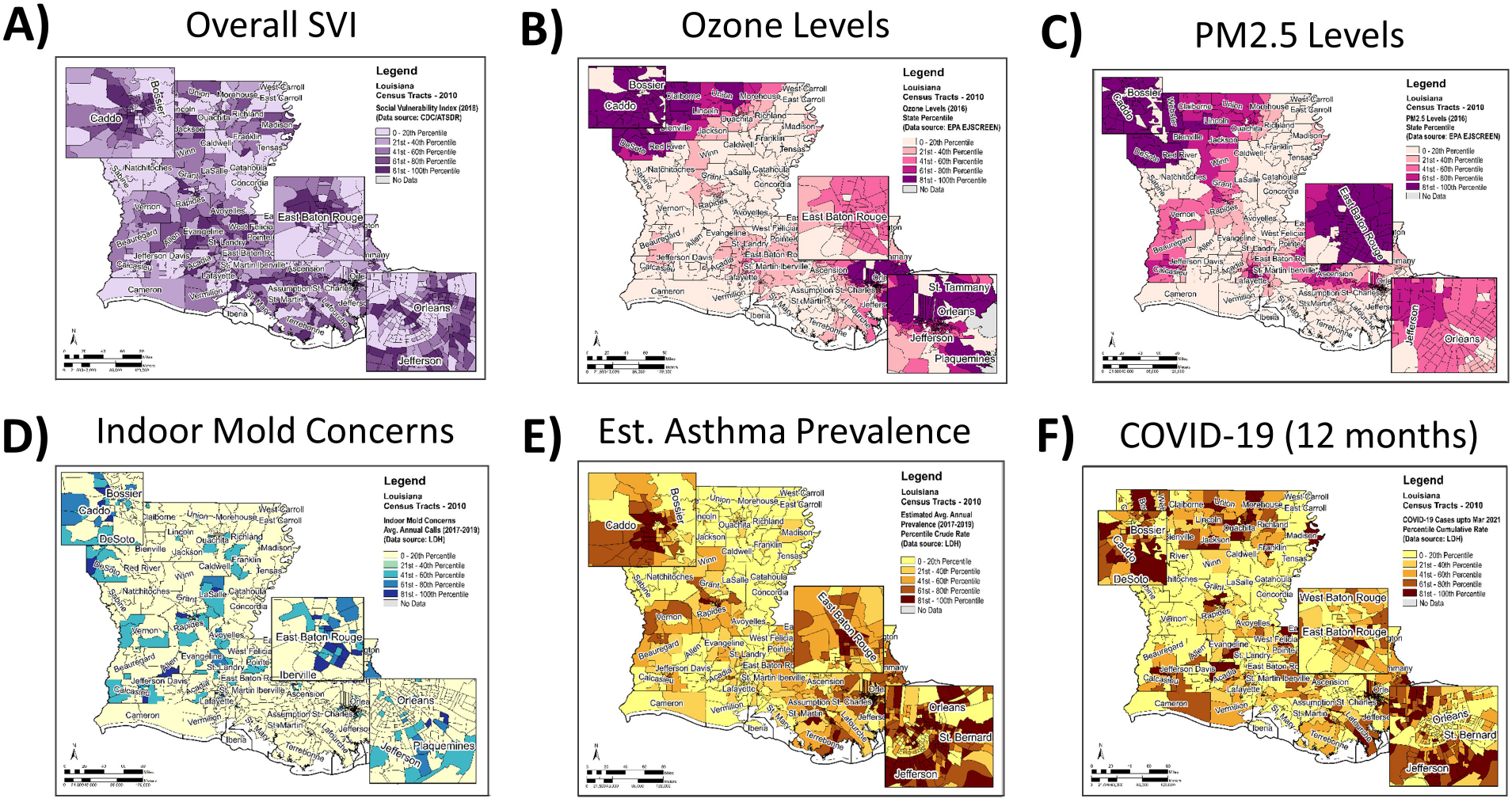
Spatial analysis of social and environmental vulnerability, asthma and COVID-19. Darker colors for each measure indicate higher percentile ranks. Most counties in LA had census tracts with SVI **(A)** above the median. Ozone **(B)** and PM_2.5_ **(C)** were higher in the northwest and southeastern parts of the state. Indoor environmental quality concerns (mostly in the form of mold complaints) were also reported from all over the state **(D)**. Estimated prevalence of asthma **(E)** was often above the median in census tracts located in the northwestern, western, southern and southeastern parishes. At the 12-month time point, cumulative COVID-19 incidence rates **(F)** were higher in the northern, northwestern and southeastern parts of the state.

Next, we attempted to identify EJ communities in Louisiana using the social and environmental vulnerability metrics. We used Overall SVI from CDC/ATSDR as the overarching metric for social vulnerability. For environmental vulnerability, we chose RHI, PM_2.5_ and Ozone levels. IEQ reports were not considered because of the sparse nature of the data. Thus, we intersected Overall SVI, RHI, PM_2.5_ and Ozone datasets **(Fig 3A)**, and defined a CT as an area of EJ concern if along with high SVI, it also had either high RHI, high PM_2.5_ or high Ozone levels. Mapping these areas revealed that the 137 CTs that fit these criteria were mostly located in the major urban centers with a few spread across the more rural areas of the state **(Fig 3B)**.

**Figure 3.**
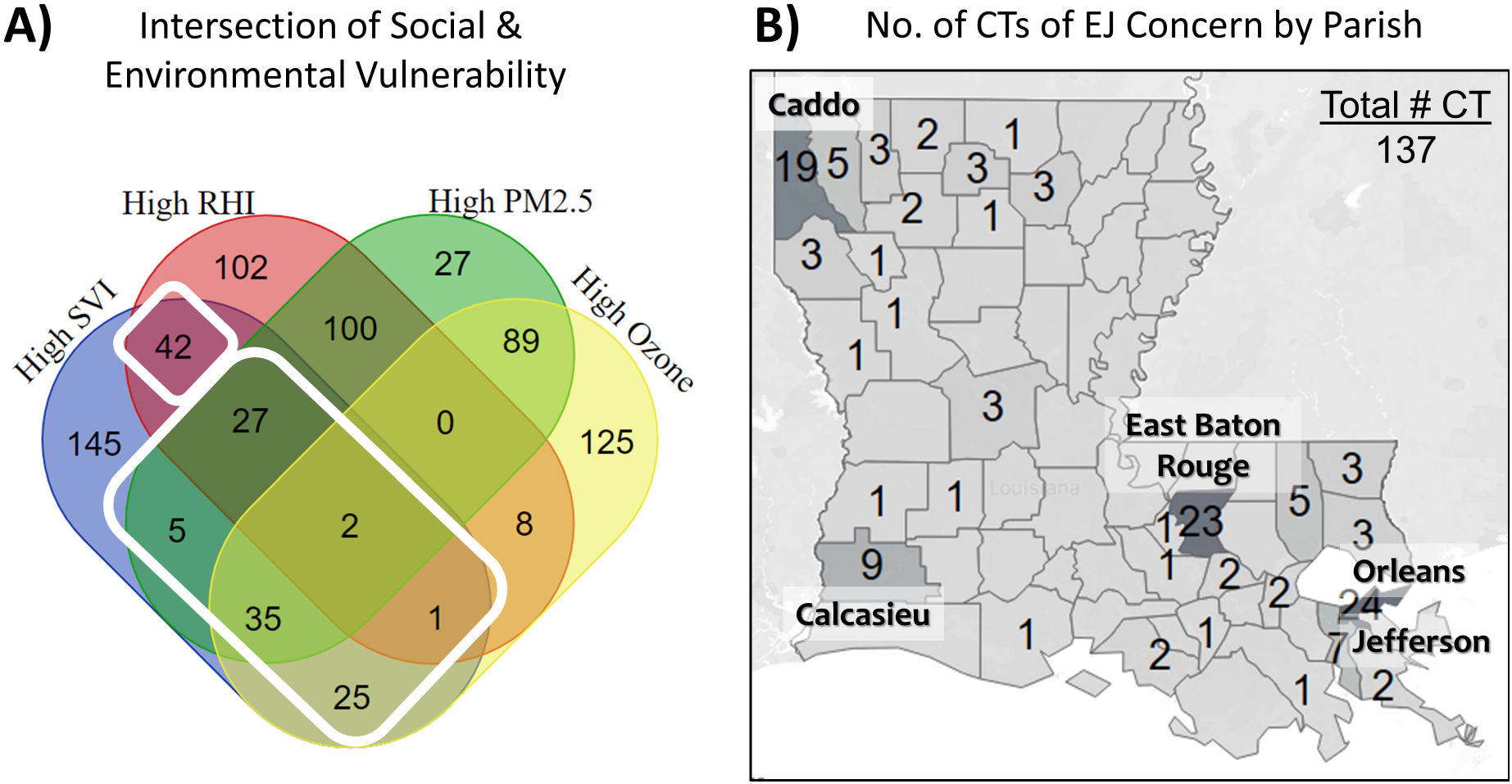
Identifying census tracts with EJ concerns. Census Tracts were defined as being of EJ concern if the Overall SVI was at or above the 75^th^ percentile, along with at least one of the environmental variables (RHI, PM_2.5_ or Ozone) at or above the 75^th^ percentile for the state **(A)**. Map shows the number of the CTs of EJ concern by Parish **(B)**. Most of them were located in the major urban areas, with a few spread across the rural areas of the state.

Finally, we surveyed the health metrics of these EJ CTs and sought to identify areas of high asthma and COVID-19 burden **(Fig 4A)**. Out of the 137 EJ CTs, 75 (55%) had high estimated asthma prevalence (at or above the 75^th^ percentile for the state), 28 (20%) had a high cumulative COVID-19 rates at the 12-month time point, and 15 CTs (11%) had a high burden of both asthma and COVID-19. The 75 EJ CTs with a high burden of asthma were mostly located in the major urban centers of the state, with a few in the more rural areas **(Fig 4B)**. A similar pattern was observed for the 15 EJ CTs with a high burden of both asthma and COVID-19, with a third of them being located in the northwestern part of the state in Caddo Parish **(Fig 4C)**.

**Figure 4.**
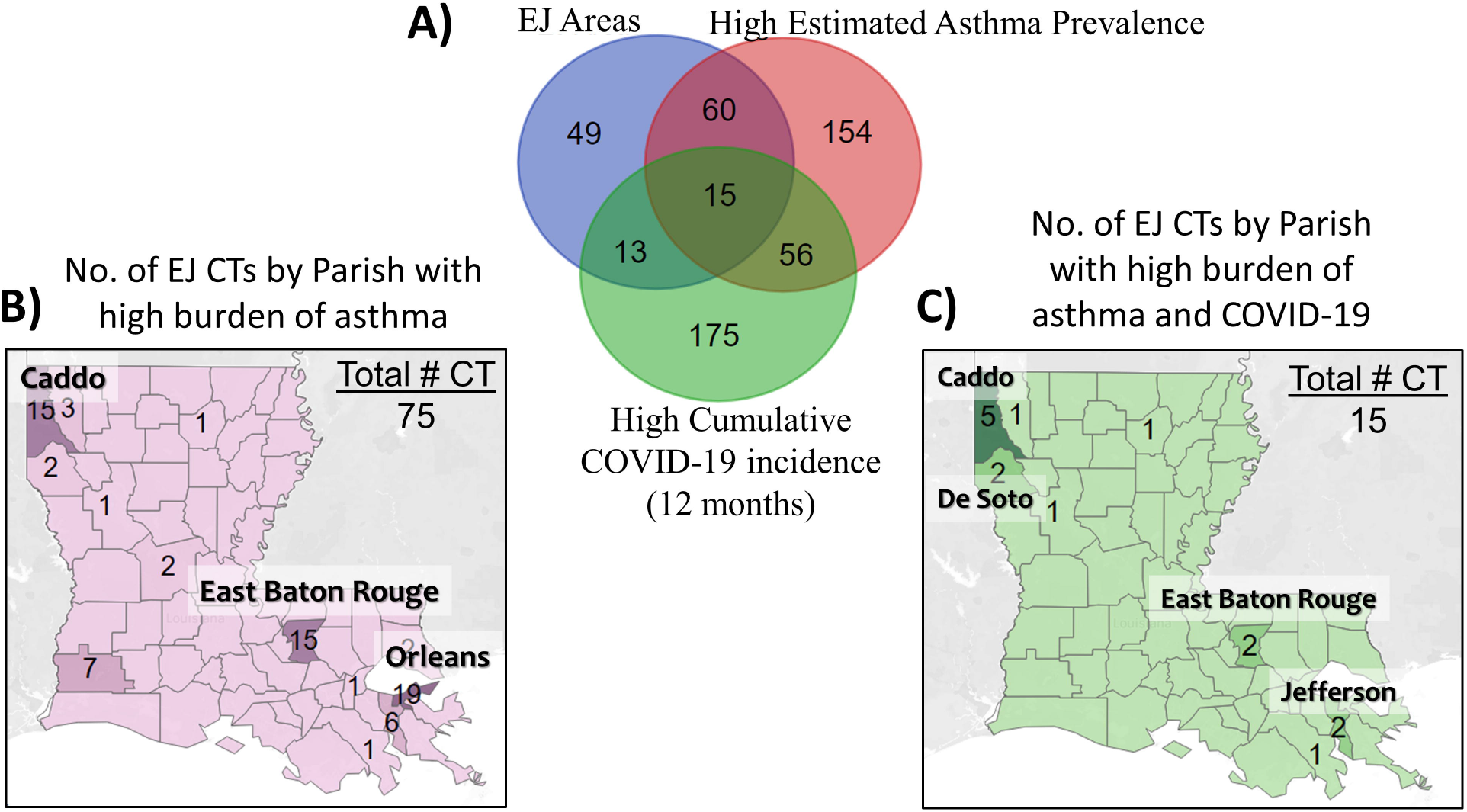
Identifying EJ communities with high COVID-19 and asthma burdens. **(A)** Of the 137 census tracts that were deemed of EJ concern, 75 (55%) had estimated prevalence of asthma at or above the 75^th^ percentile for the state. The majority of these CTs were located in the major urban areas, with others spread across the more rural areas of the state **(B)**. A similar pattern was observed for CTs that were of concern for EJ, asthma as well as COVID-19. About a third of the 15 EJ CTs deemed areas of concern for both asthma and COVID-19 were located in the northwestern part of the State in Caddo Parish **(C)**.

## Discussion

Over the past year, COVID-19 has taken a disproportionate toll on minority populations across the country, shining a spotlight on the health, socioeconomic and environmental disparities faced by them. According to the CDC, asthma is one of the underlying conditions that, when moderate to severe, may be a risk factor for severe illness with COVID-19. It is also a respiratory health condition that disproportionately affects racial and ethnic minorities [14]. Here we show that these trends in health disparities, previously assessed nationally and/or at the county level [3, 9, 14, 15], hold true at the census tract level in Louisiana. Specifically, our data show a moderately strong positive correlation between SVI due to minority status/language barrier and three health data variables: asthma hospitalization; estimated asthma prevalence; and cumulative COVID-19 incidence at 3 months **(Table 2)**. Interestingly, SVI measures were either negatively or not significantly correlated COVID-19 incidence at the 9- and 12-month time points, indicating that social vulnerability factors may have played a greater role in COVID-19 spread early in the pandemic, but may have been of diminishing importance as the pandemic wore on **(Fig 1 and Table 2)**.

Regardless, an assessment of specific social vulnerability factors, asthma and COVID-19 is likely to reveal important characteristics of minority populations that have contributed to long-standing health disparities along racial and socioeconomic lines. One such characteristic may be lack of access to a vehicle. **(Table 3)**. This may be of particular importance in Louisiana, which lacks extensive public transport infrastructure in most places, and may be a factor that prevents people from accessing areas away from their immediate vicinity to acquire healthcare services, or other commodities conducive to health (such as healthy foods, if living in a food desert). In urban areas, where public transport is available, people without vehicles may not be able to socially distance in the confined space while using public transportation. Indeed, a connection between higher subway ridership and COVID-19 infection risk was observed in a previous study in New York [16], and has been suggested previously in an earlier sub-county study in Louisiana measuring Area Deprivation Index and COVID-19 risk [17]. Thus, the possible impact of not having access to a vehicle (among other vulnerability factors) on chronic health conditions, as well as COVID-19 spread, warrants further investigation, especially in Louisiana’s minority communities that face other socioeconomic and environmental stressors as well.

Minorities, especially African-Americans, are also more likely to live in EJ communities and be exposed to environmental pollutants that can influence their health [18]. Asthma is a respiratory condition that has several known environmental triggers such as ozone and particulate matter. Various studies have also shown that there may be a connection between air pollutants affecting respiratory health (mainly, particulate matter) and COVID-19 incidence as well as mortality [9, 15, 19, 20]. In our study, however, we found higher rates of COVID-19 incidence, asthma hospitalizations and estimated asthma prevalence in CTs with higher ozone levels **(Fig 1 and Table 2)**. According to the US EPA, long-term exposure to ozone pollution can damage the lungs and make it likely to acquire respiratory tract infections [21, 22]. Ozone is also known to aggravate asthma and may be one of the factors that leads to developing asthma [23-26]. Of note, a 2018 study showed that exposure to even low levels of ozone (below the limit set by the National Ambient Air Quality Standards) had adverse effects on the respiratory health of African-American children with poorly-controlled asthma [27]. Since ground-level ozone is formed from precursors present in traffic exhaust and industrial emissions, it is possible that ozone levels may be higher in the more urban, industrialized areas of Louisiana than the more rural regions, as has been previously documented by CDC for the entire United States [28]. If this assumption holds true, it would explain our results indicating that CTs with EJ concerns are largely located in the more urban areas of the state **(Fig 3)**. Thus, any possible contribution of ozone to asthma prevalence, asthma exacerbation as well as the COVID-19 burden in Louisiana bears investigation in the areas of concern.

Since about 60% of the CTs with EJ concerns related to outdoor air also reported IEQ concerns (primarily, mold in homes), the 116 tracts identified to be of concern with regards to asthma **(Fig 4A)** may benefit from educational outreach that includes both clinical and environmental management of asthma [29-32]. To accommodate individuals with asthma who may not have access to vehicles for easy transportation, we propose either a local community-based or a home-based education model that ideally incorporates an environmental evaluation of their home. Such initiatives are not only likely to reduce respiratory health disparities in LA, but they may also open the door to further research that establish the factors underlying both COVID-19 incidence and asthma prevalence **(Table 2)**, the potential environmental impact on both health indicators, as well as any increased burden of asthma due to COVID-19 in EJ communities. Considering the fact that much of the adverse impact from COVID-19 on vulnerable populations may have been through pre-existing disparities in chronic health outcomes [1, 2], such initiatives focusing on improving health and environmental equity are likely to yield long-term gains.

Our study is the first to our knowledge that evaluates the intersection of social and environmental vulnerability, COVID-19 and asthma statewide at the census tract level in LA. Thus, it represents an important milestone in identifying and understanding specific communities that are at the crossroads of environmental justice issues as well as health disparities. However, there are some important limitations that should be considered. First, it is an ecological study, and therefore, while providing useful observational data to help generate hypotheses, it cannot be used to test any hypothesis related to population health. Second, though we chose the most current environmental vulnerability metrics available from EPA EJSCREEN, we cannot be certain that they model current environmental conditions. At the same time, much of the impact of air pollution on respiratory health is due to long-term exposure [23-27], not just short-term; therefore, it is still instructive to consider these data even if it is a few years old as of 2021. Third, there are multiple different models for estimating the burden of air pollution; all have their own pros and cons, and all are relatively more unstable at smaller geographic levels (such as sub-county geographies). The environmental vulnerability metrics and visualization scheme presented here were based on those used by a well-established resource to study environmental justice concerns (EJSCREEN). However, we cannot rule out that using different models of air pollution estimates or applying the same models but at a different spatial resolution may yield different results. Finally, due to the lack of outpatient asthma surveillance data at the sub-county level, we used inpatient discharge data to estimate the burden of asthma hospitalization as well as estimated asthma prevalence. These data are subject to various limitations that result from incomplete reporting, changes in hospital administration (such as closures or merges), inequities in healthcare access, choice of diagnosis codes, etc. Complete metadata related to asthma hospitalizations can be found through the LDH website (https://ldh.la.gov/assets/oph/Center-EH/envepi/EPHT/Infotab-Metadata/Asthma_5-1-2019.pdf).

## Conclusion

In the wake of COVID-19, there has been a spotlight on health disparities due to social and environmental vulnerability often faced by people living in EJ communities. Given that Louisiana has a high burden of both EJ and health concerns, we examined these relationships in Louisiana at the census tract level and at four different time points during the pandemic. The results show that CTs with high SVI, high PM_2.5_ and high Ozone levels had a higher burden of asthma and/or COVID-19, but the relationships only held true at certain time points **(Fig 1, Fig 2 and Table 2)**. Moreover, one of the strongest correlation coefficients was observed between COVID-19 incidence at 3 months and the estimated prevalence of asthma, indicating similar vulnerability factors may be influencing both health outcomes **(Fig 2)**. In terms of where the areas of concern were located, we identified 137 CTs, most of which were concentrated in the major urban centers of the state **(Fig 3)**. Of note, 75/137 CTs (55%) had a high burden of asthma and 15/137 (11%) had a high burden of COVID-19 as well as asthma **(Fig 4)**. Based on the results presented here, we propose that further research is warranted into the impact of COVID-19 in EJ areas, specifically as it relates to ozone exposure. Currently, much of the environmental focus of the pandemic remains on PM_2.5_ levels; however, we noted that higher levels of ozone was consistently associated with higher incidence rates of COVID-19, and it was the only environmental factor that appeared to have an additive effect over SVI on COVID-19 incidence **(Fig 1)**. Given ozone’s ability to damage lungs over time, contribute to the both development and exacerbation of asthma, as well as render people more susceptible to infections, we propose that more research is needed to elucidate the role of ozone on COVID-19 outcomes. From a public health standpoint, more programs and policies may be helpful that help people in EJ communities overcome their health, social and environmental challenges. Examples include those that assist with improved access to healthcare, transportation, healthy foods, better employment opportunities, healthy housing, air quality monitoring, and environmental health education, among others. Considering the adverse impact of the pandemic on vulnerable communities facing long-standing disparities in health and environment, programs such as these (and further data that support their efficacy) are likely to be important not only for improving health equity but also future pandemic preparedness for the entire nation.

## Data Availability

Social Vulnerability, Environmental and COVID-19 data are publicly available, and links are provided in the manuscript. The Asthma and Indoor Environmental Quality data cannot be posted publicly due to HIPAA restrictions, but can be made available by LDH upon request, following appropriate suppressions and IRB reviews as necessary.

## Acknowledgements

We extend our sincere thanks to the LDH Section of Infectious Disease Epidemiology and Bureau of Health Informatics for their tireless efforts and dedication at gathering and publishing reliable health data for the State of Louisiana, while simultaneously tackling the COVID-19 pandemic.

